# PRS-GRID: A Cross and Within Ancestry Polygenic Risk Prediction Method Based on Individual Genetic Distance

**DOI:** 10.1101/2024.05.16.24307490

**Authors:** Liwei Tang, Cong You, Xue-jun Kong, Valerio Napolioni, Jie Huang

**Affiliations:** Department of School of Public Health and Emergency Medicine, Southern University of Science and Technology, Shenzhen, China; Beijing International Center for Mathematical Research, Peking University, Beijing, China; SYNAPSE Center, Athinoula A. Martinos Center for Biomedical Imaging, Massachusetts General Hospital, Charlestown, MA, USA; Genomic and Molecular Epidemiology (GAME) Lab., School of Biosciences and Veterinary Medicine, University of Camerino, Camerino, Italy; Institute for Global Health and Development, Peking University, Peking, China

**Keywords:** Polygenic risk score, multiple ancestry, genetic distance

## Abstract

**Background:** Two decades of genome-wide association studies (GWAS) have led to the fast-growing application of polygenic risk prediction (PRS). However, due to population structure and evolutionary path differences, the PRS substrate derived mostly from studies of European ancestry does not work equally well for other ancestries. There is an association between prediction accuracy decay and individual genetic distance (GD) to the genetic centers (GC) of various populations.

**Objectives:** To develop a new PRS method and software that utilizes individual GD to improve PRS risk prediction accuracy, especially for non-European populations.

**Method:** We hypothesize that adding a GD-based weight into PRS methods would enhance its risk prediction performance, particularly for minority groups. We explore the GD first by principal components (PC) and then by phylogenetic tree structures. Building on top of an emerging software (PRS-CSx) that achieves high prediction accuracy across multiple-ancestries, we present PGS-GRID, where “GRID” stands for “**<u>G</u>**enetic **<u>R</u>**eference based on **<u>I</u>**ndividual **<u>D</u>**istance”.

**Results:** We developed a preliminary version of PRS-GRID and pilot tested its prediction performance for a classic quantitative trait (*e*.*g*., height) and a disease trait (*e*.*g*., type-2 diabetes). We found slight but noticeable improvement of risk prediction in minority populations. We further explored a random forest approach so that the performance of PRS-GRID could be clearly explained, which is a key step for PRS to be used in clinical and public health practice.

**Conclusions:** The PRS-GRID philosophy and method represent an innovative and significant advancement in the field of polygenic risk prediction. Our work provides a foundation for future research and clinical applications aimed at reducing health disparities and improving population health through personalized medicine.

## Introduction

Two decades of genome-wide association studies (GWAS) have led to substantial progress in discovering an increasing number of alleles contributing to the risk of complex traits. This largely conforms to the ‘common variant–common disease’ hypothesis.^1^ A straight-forward application of GWAS is polygenic risk prediction (PRS). Early PRS analyses focused on variants that fell below the genome-wide significance threshold (*P* < 5 × 10^−8^).^2^ However, the inclusion of less significant genetic variants has been found to improve PRS prediction.

A landmark study in 2009 demonstrated the theoretical rigor and technical feasibility of including thousands of genomic variants that are not genome-wide significant for risk prediction.^3^ Almost a decade later, another study showed that a PRS constructed from millions of SNPs could predict the risk for cardiovascular diseases equivalent to monogenic mutations.^4^ Since then, it has become common practice to use thousands and even millions of genetic variants to derive genetic risk predictions for complex traits. Several studies have shown that PRS constructed in such a manner can be more predictive than well-established traditional clinical risk factors.^5^ As the number of genetic variants increases, the inherent issue related to genetic linkage disequilibrium (LD) needs to be properly tackled. Advances in computational methods have created software tools that adjust or shrink the effect size (β value) of linked genetic variants.^6-9^ Now, it is not unusual for large GWAS consortia to release adjusted or shrunk PRS reference panels in the public domain, such as the PGS Catalog.^10^

For both quantitative traits and binary disease traits, a key stake is to enable and improve risk prediction across diverse ancestries to reduce the inequity of PRS in minority populations. Recently, large GWAS consortia have begun to release multiple-ancestry GWAS and PRS references for both quantitative traits (*e*.*g*., height)^11^ and disease traits (*e*.*g*., type-2 diabetes, T2DM).^12^ In 2022, PRS-CSx^13^ emerged and has been shown to outperform many other PRS tools that aim to improve genetic risk prediction accuracy across multiple ancestries. The current PRS-CSx software can be run in two ways. The simpler way is utilizing the ‘meta’ option to produce one set of SNP weights that can be applied to all ancestral groups and all individuals, while the preferred way is to learn a linear combination of population-specific PRS for each ancestral group. The former does not need a validation set to learn the linear combination weights, while the latter approach requires assigning target samples into distinct ancestral groups and learning the linear combination weights in the validation set, which are then applied to the testing set.

RPS-CSx assigns individuals into ancestral groups without fully considering each individual’s genetic characteristics. A 2023 study pointed out that genetic accuracy decays not only across different ancestries but also within populations^14^. It used principal components (PC) as a metric to evaluate the PRS accuracy decaying problem. Some other studies used Uniform Manifold Approximation and Projection (UMAP) to characterize genetic ancestry.^15^ However, genetic distance (GD) was only used as a measurement of the problem (reduced prediction accuracy) and not as a potential solution. We hypothesize that adding a GD-based weight into the regression step of PRS-CSx would further enhance its risk prediction performance, especially for minority groups.

In this study, we calculate and incorporate genetic distance at the individual level and explore it as a potential solution to improve PRS-based risk prediction across multiple ancestries. We explore the genetic distance first by PC and then by phylogenetic tree. It has been reported that complex traits are strongly associated with evolutionary development and positive selection.^16^ Using objectively derived genetic ancestry instead of self-reported race would improve PRS for common diseases such as coronary artery diseases. Building on top of PRS-CSx,^13^ we present PGS-GRID, where “GRID” stands for “**<u>G</u>**enetic **<u>R</u>**eference based on **<u>I</u>**ndividual **<u>D</u>**istance”. We test and evaluate the prediction performance of PRS-GRID for a classic quantitative trait (height)^11^ and a disease trait (T2DM).^12^

## Method

PRS-GRID is designed to generate multi-ancestry PRSs by incorporating individual GD from the genetic center (GC) of representative populations, including those with smaller GWASs. The method has four key steps (**Fig. 1**): (**1**) Construct PRS reference for each ancestry using PRS-CSx; (**2**) Determine the GC of reference populations and calculate GD for each individual to those GCs; (**3**) Train the association between the trait of interest and PRS by incorporating each individual’s GD; (**4**) Validate the trained parameters in the validation dataset and evaluate the model’s prediction accuracy for both quantitative traits and binary disease traits.

**Fig 1.**
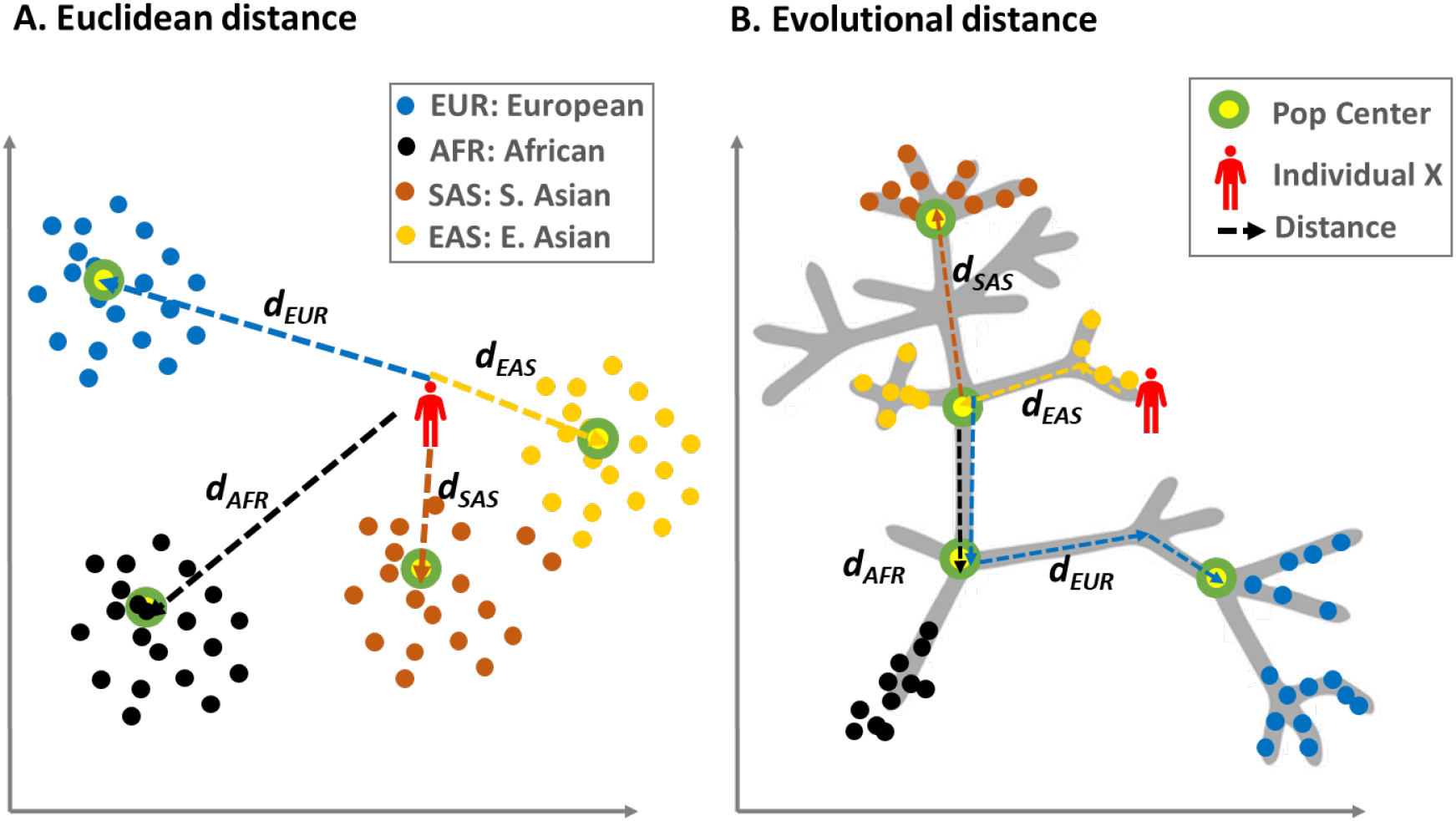
Individual distance in PRS-GRID. ^*^ Colorful dots indicate subjects of distinct populations. Green circles with yellow background indicate the center of each population. The dotted lines indicate distance from a target person (red) to the center of each population. Panel **B** uses Darwin’s “Tree of Life” as an illustrative pedigree. The center of each population is at the root of each sub-pedigree, and the distance is measured along the tree branches.

### Study Participants

We used data from the UK Biobank, an ongoing prospective cohort study designed to investigate the determinants of diseases in middle and older age.^17^ Participants were recruited between 2006-2010 and completed a comprehensive assessment of health and lifestyle including a touchscreen questionnaire, interview, measures of physical function and biological samples.^18^ The study population comprised individuals from the UK Biobank who self-reported their race as white, black, Asian, or Chinese. We determined the ancestral groups of the study population by comparing self-reported races with the results of PC analysis on ancestral groups in accordance with criteria suggested by previous studies.

### Assessment of GD

We first extracted each person’s PC values provided by the UK Biobank data (data field: 22009). We then used the median values of PCs as the GC of four ethnic groups: European (EUR), African (AFR), Southern Asian (SAS), and Eastern Asian (EAS). We then calculated the Euclidean GD of each individual to these four centers as illustrated by Ding *et. al*.^14^ We included all 40 PCs for this analysis. We next used yhaplo (https://github.com/23andMe/yhaplo), which can identify the Y-chromosome haplogroup of each male in a sample with up to millions of individuals. This method first builds an internal representation of the Y-chromosome phylogeny by reading its primary structure from (Newick-formatted) text and importing phylogenetically informative SNPs from the ISOGG database. It then affiliates each SNP with the appropriate node and grows the tree as necessary. It traverses the tree for each individual, identifying the path of derived alleles leading to a haplogroup designation.

### Train PRS Model with Individual GD

The raw Euclidean distance (dE) of the GD for each individual is given in **Equation 1**. PC_1_ to PC_n_ are the PCs of each individual, while PC_c1_ to PC_cn_ are the presumed genetic centers of each race. The GD is then normalized through **Equation 2**, where d_1_ to d_4_ are the GD from four main races or ancestral groups (EUR, AFR, SAS, and EAS). β_adj,r_ is the adjustment coefficient and r belongs to (EUR, AFR, SAS, EAS).

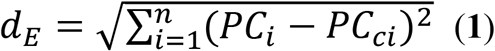

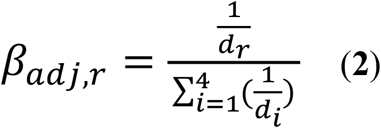

We fit the target phenotype and PRS with the GD adjusted coefficient as shown in **Equation 3**, where PRS_GRID_ is the adjusted PRS score given by PRS-GRID and the PRS_CSx_ is the PRS score given by PRS-CSx.

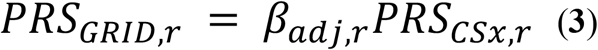

### Validate PRS in an Independent Set

Linear regression (**Equation 4**) and logistic regression (**Equation 5**) are used to fit for the prediction of height and T2DM outcome, respectively. X_i_ is the ith PRS and β_i_ is the regression coefficient for the independent variable X_i_ given by best-fitting logistic regression models. The X_c_ and β_c_ correspond to the covariate and the regression coefficient of the covariate.

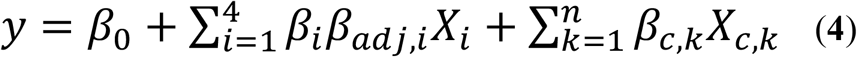

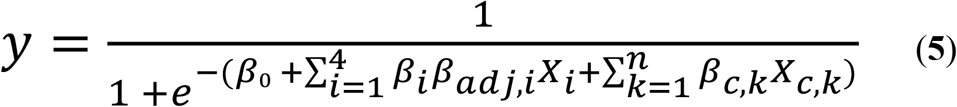

### Evaluate Prediction Performance

For providing robust results of the model performance, we used 10-fold cross-validation in our analyses. The R-squared and AUC (Area Under the Curve) were respectively used to evaluate the predictive performance for the continuous numerous values of height and the binary outcome of T2DM. To compare the impact of introducing correction coefficients on the predictive power of each variable in the model, we utilized the random forest approach for analysis and visualization. In the random forest approach, the percentage increase in mean squared error (% IncMSE) and the mean decrease accuracy reflect the importance of each variable in the prediction model. The age and sex of the participants were included in the regression models as the covariates to adjust for the possible confounding effects.

## Results

After excluding individuals lacking phenotype data (T2DM binary outcome and height) and necessary covariates (age and sex), we included a total of 473,399 and 472,123 individuals for the prediction of T2DM and height, respectively, using ancestral-specific PRS scores from PRS-CSx. The ancestral groups and the outcome phenotypes of the study population are detailed in **Table 1**.

**Table 1.** Baseline characteristics of the study population.

| Characteristic | EUR | AFR | SAS | EAS |
| --- | --- | --- | --- | --- |
| Age (year) | 56.8 (8.02) | 51.9 (8.05) | 53.4 (8.47) | 52.3 (7.64) |
| Female (%) | 45.72 | 42.61 | 53.3 | 37.22 |
| **Alcohol status** |  |  |  |  |
| Never | 14,646 | 1,147 | 3,214 | 339 |
| Previous | 15,951 | 389 | 419 | 53 |
| Current | 425,989 | 5,494 | 4,187 | 1,044 |

| Smoking status |  |  |  |  |
| --- | --- | --- | --- | --- |
| Never | 245,732 | 4,973 | 6,098 | 1,145 |
| Previous | 162,102 | 1,190 | 998 | 189 |
| Current | 47,532 | 848 | 711 | 102 |
| Height(cm) | 168.67 (9.25) | 167.36 (8.65) | 164 (9.25) | 161.62 (7.83) |
| T2DM |  |  |  |  |
| Case | 34,198 | 1,234 | 1,917 | 103 |
| Percentage % | 6.98 | 15.04 | 19.89 | 6.71 |

### Prediction Performance

We selected the T2DM and the height of the individuals from the UK Biobank as the phenotype variables to examine the prediction accuracy of our PRS-GRID approach. The prediction performance and the comparison between the original PRS-CSx and ancestral genetic distance adjusted PRS-GRID models were as shown in **Figure 2**. For T2DM, the AUC of all three underrepresented ancestral groups were slightly higher than the original PRS-CSx models by 10-fold cross validation. The AUC of the ancestral groups of AFR, SAS, and EAS were 0.697, 0.819, and 0.733 in the original PRS-CSx model, while 0.717, 0.826, and 0.753 in the PRS-GRID model, respectively.

**Fig 2.**
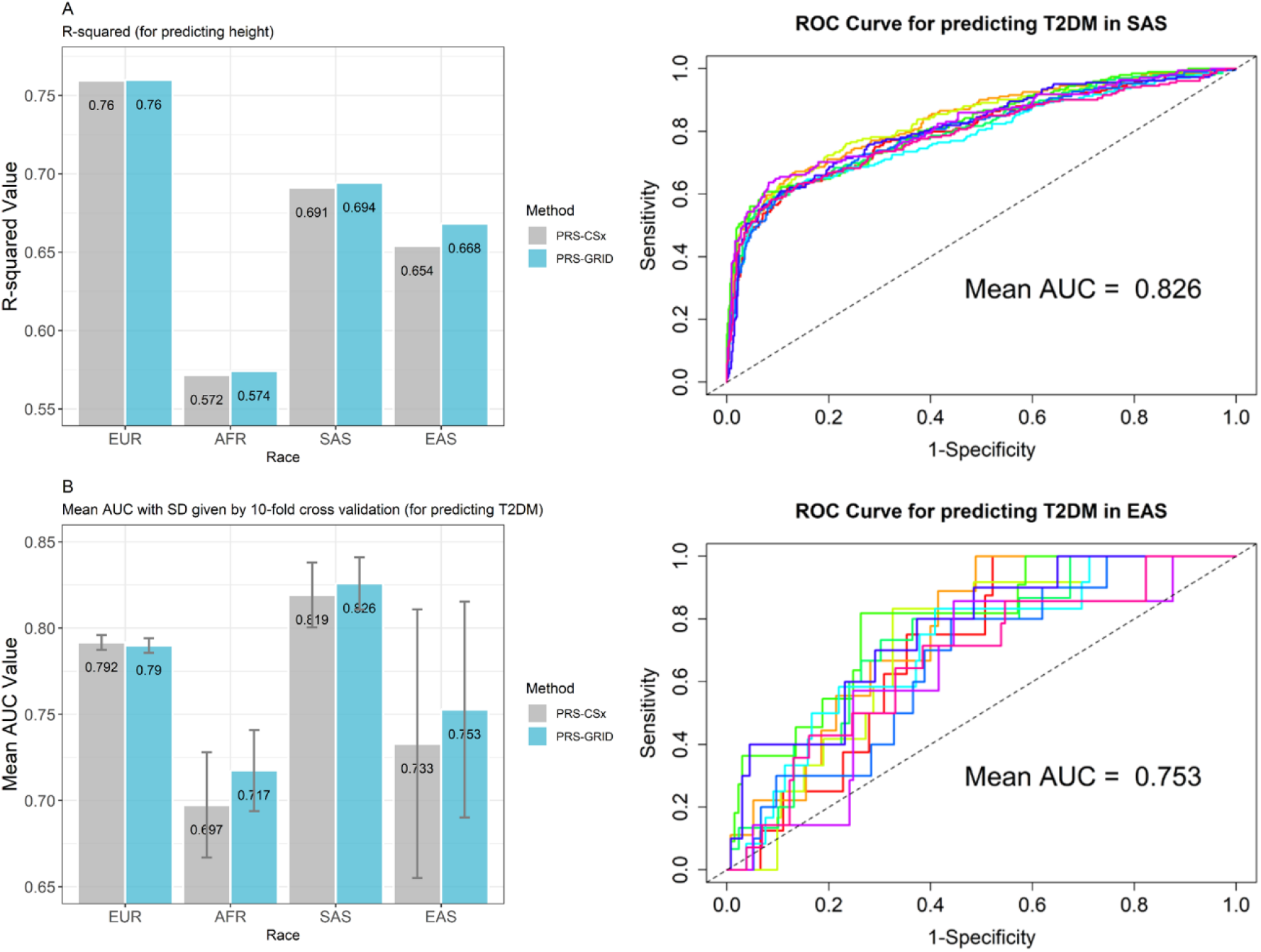
The prediction accuracy PRS-CSx vs. PRS-GRID.

### Explainable Machine Learning

To better investigate and explain the improvement on the prediction performance of PRS-GRID, we used random forest approach, a classical machine learning approach based on decision tree algorithm to explain the potential impact of the PRS scores after adjusting for the GD of multi-ancestry. **Figure 3** displays the mean decrease accuracy of four ancestral-specific polygenic risk score (PRS) models for predicting binary T2DM classification among SAS and EAS individuals, using the best-fitting random forest model. After adjusting for GD within the four ancestral GC, there was a marked improvement in the predictive accuracy of the ancestral-specific PRS models, particularly among EAS individuals. This finding supports the hypothesis that improving prediction performance after GD adjustment is due to an increase in the PRS scores from the same ancestral groups as the population being predicted.

**Fig 3.**
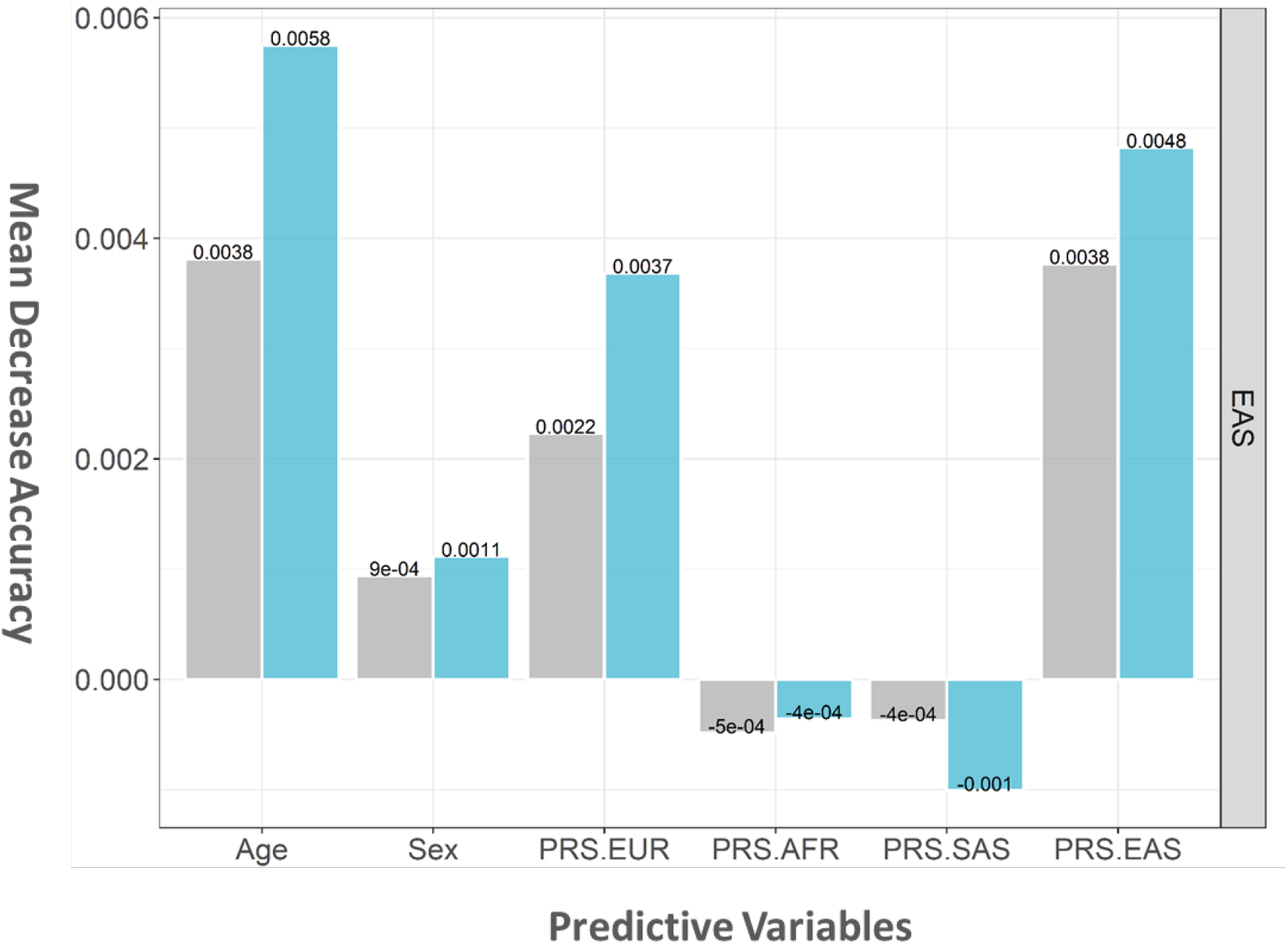
Explainable machine learning

## Discussion

Based on a recent observation that PRS prediction accuracy decays with GD from each individual to the center of genetic ancestry groups, we hypothesized that adding GD into PRS models would increase prediction performance especially for individuals of minority ancestry groups. We derived ancestry-specific reference PRS from an existing method (PRS-CSx) but mitigated its drawback of not adequately considering the heterogeneity of individuals within the same ancestral group.

The lofty objective of PRS is precision medicine, where an individual’s genetic profile is the key. To fully take into account each individual’s characteristics, we measure each person’s genetic position on the global population map by calculating its genetic distance to the center of reference populations. The genetic distance-based weight adjustment in the PRS-GRID model provides a novel way to enhance the utility and accuracy of PRS across different ancestries. This method can potentially reduce health disparities by providing more accurate genetic risk assessments for diverse populations. Our findings suggest that incorporating genetic distance in PRS calculations can lead to more equitable healthcare outcomes by ensuring that PRS models are applicable to all ancestral groups, not just those of European descent.

We developed and tested PGS-GRID, a new method to improve cross-ancestry PRS prediction by incorporating individual GD. Our results demonstrated that the PRS-GRID model performed better than traditional PRS models and PRS-CSx models in predicting both height and T2DM across different ancestries. The improved prediction performance of the PRS-GRID model highlights the importance of considering genetic distance in polygenic risk prediction models. Our study confirms that accounting for individual genetic characteristics can mitigate the decay in prediction accuracy when applying PRS models across diverse populations. This approach is particularly beneficial for minority populations, who often have limited representation in GWAS and PRS studies.

In addition to the improved predictive accuracy, the PRS-GRID model offers several practical advantages. By using individual genetic distances rather than relying solely on population-specific adjustments, the PRS-GRID approach allows for a more personalized risk assessment. This is particularly relevant in increasingly diverse societies where individuals may have mixed ancestry and traditional PRS models may not accurately capture their genetic risk. Moreover, the PRS-GRID model’s framework can be adapted to other complex traits and diseases, making it a versatile tool for genetic epidemiology. Future research should focus on validating the PRS-GRID model in larger and more diverse cohorts to ensure its robustness and generalizability. Additionally, integrating other omics data such as epigenomics and transcriptomics, could further enhance the predictive power of the PRS-GRID model.

For PRS to be eventually usable in clinical practice, the explainability is key. By comparing the mean decrease in accuracy of the four ancestral-specific PRS scores before and after adjusting for the genetic distance adjustment coefficients, it can be partly explained whether and how the introduction of genetic distance-based adjustment coefficient influences the prediction ability of the models.^19^ The implementation of PRS-GRID in clinical practice could revolutionize the way genetic risk is assessed and managed. By providing more accurate risk predictions, healthcare providers can tailor prevention and intervention strategies to individuals’ specific genetic profiles, leading to better health outcomes. However, the transition from research to clinical application will require careful consideration of ethical, legal, and social implications, including issues related to genetic privacy and data sharing.

The present study still has some limitations that should be acknowledged. First, although we made an effort to improve the prediction performance of PRS scores across populations by adjusting for multi-ancestral GD, the optimal measurement of GD in populations with uncertain population structure requires further exploration. Our next-step studies will consider combining results from phylogenetic tree analysis and molecular anthropology to develop more effective assessment methods for multi-ancestral GD. Second, the sample size of the minority group, specifically individuals of EAS, in the UK Biobank used for this study was small. The limited sample size may compromise accurate assessments of the predictive performance of the models among these underrepresented ancestral groups. Additionally, it is important to consider that many human diseases are influenced by multiple complex factors, including genes, environmental exposure, and gene-gene or gene-environment interactions. Our study did not include these multiple factors due to the lack of data on gene-environment interactions in the UK Biobank dataset. However, we plan to incorporate this information in future studies once sufficient data becomes available.

## Conclusions

The PRS-GRID model represents an innovative and significant advancement in the field of polygenic risk prediction. By incorporating individual genetic distance, it addresses key limitations of traditional PRS models and offers a more accurate and equitable approach to genetic risk assessment. Our study provides a foundation for future research and clinical applications aimed at reducing health disparities and improving population health through personalized medicine.

## Data Availability

All data produced in the present study are available upon reasonable request to the authors

## Author contributions

All authors meet authorship criteria by contributing to components of research conception, design, interpretation of results, and manuscript revisions. JH. conceptualized and designed the study, conducted formal data analyses, and drafted the initial manuscript. L.T., C.Y. conducted formal data analyses, interpreted the results, and revised the manuscript. X.K., V.N. provided input into the study design and revised the manuscript. All authors reviewed the final manuscript as submitted.

## Acknowledgments

This research was conducted using the UK Biobank resources under application 66137. We thank the participants for sharing their health-related information.

## Ethical statement

All participants provided written informed consent at their baseline visit. Ethical approval of the UK Biobank study was obtained from the National Information Governance Board for Health and Social Care and the National Health Service North West Multi-Centre Research Ethics Committee (Ref 11/NW/0382).

